# Risk factors associated with mortality in critically ill patients with SARS-CoV-2 pneumonia in a tertiary hospital in Ecuador

**DOI:** 10.1101/2025.05.15.25327696

**Authors:** Fausto Guerrero-Toapanta, Jorge Hurtado-Tapia, Abel Godoy-Miketta, Yeimi Herrera-Parra, Freddy Maldonado-Cando, Gabriel García-Montalvo, José Vinueza-Rivadeneira, Juan López-Altamirano, Edison Ramos-Tituaña, Cecilia Cruz-Betancourt

**Author notes:** Correspondence Author: Dr. Fausto Guerrero-Toapanta, MSc., Universidad de las Américas (UDLA), Vía a Nayón, Quito 170124.

## Abstract

**Objectives:** SARS-CoV-2 pneumonia in critically ill patients is characterized by a high mortality rate. Numerous risk factors have been identified for this condition. The objective of this study was to determine the risk factors for mortality in critically ill patients with SARS-CoV-2 pneumonia receiving invasive mechanical ventilation.

**Methods:** This was an observational and retrospective study of patients who were in the intensive care unit of a third-level hospital between March 2020 and December 2021. Adults with confirmed SARS-CoV-2 infection and who were in the unit for more than 48 hours were included. Demographic, clinical, laboratory, and mechanical ventilation data were collected. All patients received protocolized treatments. Univariate and multivariate analyses were performed, with a p value of < 0.05, and the R statistical tool was used.

**Results:** Of 1024 patients, 203 were analyzed. In the univariate analysis, the age, weight, hypertension status, Simplified Acute Physiology Score III, Charlson Comorbidity Index score, neutrophil/lymphocyte ratio and acute respiratory distress syndrome status significantly differed between the living and deceased patients groups. In the multivariate analysis, the Charlson Comorbidity Index score (OR 1.36, 95% CI 1.12-1.68, p=0.002), acute respiratory distress syndrome (OR 4.16, 95% CI 1.24-16.04, p=0.02), and neutrophil/lymphocyte ratio (OR 1.040, 95% CI 1.00-1.07, p= 0.02) remained statistically significant. The mortality rate in the unit was 58.1%.

**Conclusions:** The Charlson Comorbidity Index score, neutrophil/lymphocyte ratio and acute respiratory distress syndrome status were associated with increased mortality.

## INTRODUCTION

SARS-CoV-2 causes COVID-19, and patients with severe pneumonia requiring admission to the intensive care unit (ICU) have high mortality rates in developing countries ^1,2^. Clinical-epidemiological suspicion and prompt and adequate treatment influence the mortality of these patients. Septic shock and acute respiratory distress syndrome (ARDS) can complicate the condition of patients with severe pneumonia ^3,4^ and increase patient mortality.

During the COVID-19 pandemic, an extensive list of mortality risk factors was identified in patients with severe COVID-19 ^5–11^. To date, the number of publications concerning the mortality risk factors in patients with severe SARS-CoV-2 pneumonia admitted to the ICU in our country remains limited ^12–15^.

The main objective of this study was to determine the risk factors for mortality in critically ill patients with SARS-CoV-2 pneumonia receiving invasive mechanical ventilation (IMV) who were admitted to the ICU of a third-level hospital in the city of Quito-Ecuador from 2020 to 2021. The secondary objectives were to characterize the patients demographically and clinically, establish the frequencies of sepsis and ARDS, and determine patient mortality.

## MATERIAL AND METHODS

Study design and context: The study was approved by the Ethics Committee on Human Research of the hospital with the code IESS-HCAM-CEISH-2022-0012. An observational retrospective analytical study was carried out with patients admitted to a 35-bed medical-surgical ICU, a unit that, in 2023, had 1,685 admissions with an average stay of 6 days, 17% mortality, and 91% bed occupancy and cared for patients with an average age of 55 years, whose most prevalent pathologies were sepsis and acute respiratory failure. This ICU receives patients with clinical and surgical pathologies, such as postoperative care after highly complex surgery, transplants, etc., at the Carlos Andrade Marín Specialty Hospital (HECAM), a 500-bed hospital that belongs to the Ecuadorian Social Security Institute (IESS), in the city of Quito-Ecuador, located 2,800 meters above sea level.

Participants: The study’s sample population comprised all critically ill patients who had been diagnosed with pneumonia caused by SARS-CoV-2 from March 28, 2020, to September 18, 2021. The inclusion criteria were patients over 18 years of age admitted to the ICU with a diagnosis of severe pneumonia confirmed by SARS-CoV-2 real-time polymerase chain reaction (RT-PCR) via nasal swab or tracheal aspirate, who received IMV from the first 24 hours of admission and who remained in the unit for more than 48 hours. The exclusion criteria were pregnant patients, patients who were hospitalized for more than 48 hours before ICU admission, patients who were intubated for more than 48 hours before ICU admission, and patients with insufficient data in their records.

Data from the first 24 hours of entry were collected from electronic medical records, considering only the worst data when repeated metrics were present. Demographic and clinical variables included age, sex, weight, height, etc.; underlying diseases included obesity, diabetes mellitus, arterial hypertension, congestive heart failure, chronic kidney disease with renal replacement therapy, chronic obstructive pulmonary disease, diffuse interstitial lung disease, and immunocompromised status; severity scores included those from the Charlson Comorbidity Index, Acute Physiology And Chronic Health Evaluation II (APACHE II), Simplified Acute Physiology Score III (SAPS III), and Sequential Organ Failure Assessment (SOFA); complications included the presence of acute respiratory distress syndrome (ARDS), sepsis, septic shock, and acute kidney injury; treatments included steroids (dexamethasone or methylprednisolone), vasoconstrictor support (norepinephrine and/or adrenaline), and intravenous antimicrobials; laboratory variables included lactate dehydrogenase (LDH), creatine phosphokinase (CPK), procalcitonin, leukocytes, neutrophils, lymphocytes, the neutrophil/lymphocyte ratio, D-dimers, interleukin 6, ferritin, C-reactive protein, creatinine, total bilirubin, and mean platelet volume; respiratory variables included the ratio of partial pressure of arterial oxygen divided by fraction of inspired oxygen (PaO_2_/FiO_2_); ventilatory and pulmonary monitoring parameters included positive end-expiratory pressure (PEEP), compliance, plateau pressure, driving pressure, expiratory resistance, prone position, ratio of tidal volume divided by predicted weight (VT/kg), and recruitment maneuvers; and outcome variables included the duration of mechanical ventilation, length of stay in ICU, total length of stay, ICU outcome, and outcome at 28 days. In the case of ventilatory variables, respiratory therapy notes were also searched. All patients received management and treatment based on institutional protocols for severe community-acquired pneumonia (CP), COVID-19, sepsis, and ARDS. The data collection instrument was a spreadsheet based on the study variables, with coding and anonymization of the data.

Definitions: SARS-CoV-2 detection was performed by real-time polymerase chain reaction (RT-PCR). This test detects the nonstructural ORF1 a/b region and the nucleocapsid (N) structural protein-encoding gene unique to the virus. This process uses internal controls, which are intended to monitor proper processing in the sample purification and nucleic acid amplification steps, as well as to control the presence of RT-PCR inhibitors. NC was defined based on the criteria of the Infectious Diseases Society of America (IDSA) ^16^. The severity of CP was defined based on the criteria of the American Thoracic Society (ATS) ^17^(18) and the BERLIN consensus for ARDS ^18^. The primary outcome was the determination of mortality risk factors, and the secondary outcomes were the frequencies of ARDS and sepsis and patient mortality in the ICU.

Statistical analysis: Sample size calculations were not performed because all patients meeting the inclusion criteria were included. For descriptive statistics, categorical variables are reported as percentages, and continuous variables are expressed as medians with interquartile ranges [25-75]. To establish the risk factors for mortality, two groups were defined: living and deceased. To determine the variables with the greatest impact on mortality, in the case of numerical variables, Student’s t tests were used, according to the central limit theorem, to establish differences at 5% significance between the mortality and survival groups, and chi-square tests were used at the same level of significance for categorical variables, thus reducing the number of variables, choosing a total of 11 that showed significant differences. Using these variables, a logistic regression model was constructed whose dependent variable was mortality, and according to the Z statistic at 5% significance, 5 variables contributed significantly to the proposed mortality model. To validate the obtained model, indicators such as the area under the ROC curve and the confusion matrix were used to determine the classification of the model on mortality cases. The information of the present study was analyzed with the statistical package R (version 4.0).

## RESULTS

### Participants

During the study period, 1024 patients were admitted to the unit. From this potentially eligible group, 203 patients who met the inclusion and exclusion criteria were selected. All 203 patients underwent statistical analysis. Thus, a group of subjects was obtained with median characteristics (interquartile range [Q1-Q3]) of 62 years [52-70] and 72 kilos [65-81], 162 patients were male (79.80%), 56 patients had a body mass index greater than 30 (27.58%), 35 patients had a history of diabetes mellitus (17.24%), 73 patients had a history of arterial hypertension (35.96%), 2 patients had a history of diffuse interstitial disease (0.98%), 5 patients had chronic obstructive pulmonary disease (2.46%), 7 patients had congestive heart failure (3.44%), 11 patients had chronic kidney disease on renal replacement therapy (5.41%), and 10 patients were immunocompromised (4.92%). The median severity scores (interquartile range [Q1-Q3]) were 23 [17-25] from APACHE II, 63.5 [53-73] from SAPS III, 9 [7-12] from SOFA and 2 [0-3] from the Charlson Comorbidity Index. The demographic and clinical characteristics between the groups of living and deceased patients are shown in Table 1.

**Table 1.** Demographic and clinical variables.

| **Characteristics** | **Pneumonia due to SARS-CoV-2, living (n=85)** | **Pneumonia due to SARS-CoV-2, deceased (n=118)** | **P<0.05** |
| --- | --- | --- | --- |
| Age, years, median [Q1-Q3] | 57 [49-65] | 66 [49-65] | <0.001 |
| Male gender % | 77.64 (66/85) | 81.35 (96/118) | 0.516 |
| Weight, kilos, median [Q1-Q3] | 75 [67-85] | 70 [65-80] | 0.023 |
| Body mass index greater than 30, % | 34.11 (29/85) | 22.88 (27/118) | 0.860 |
| Diabetes mellitus, % | 14.11 (12/85) | 19.49 (23/118) | 0.317 |
| Hypertension, % | 27.05 (23/85) | 42.37 (50/118) | 0.024 |
| Diffuse interstitial lung disease, % | 0 (0/85) | 1.69 (2/118) | 0.227 |
| Chronic obstructive pulmonary disease, % | 1.17 (1/85) | 3.39 (4/118) | 0.315 |
| Congestive heart failure, % | 1.17 (1/85) | 5.08 (6/118) | 0.132 |
| Chronic kidney disease with renal replacement Therapy, % | 2.35 (2/85) | 7.62 (9/118) | 0.101 |
| Immunosuppression, % | 3.52 (3/85) | 5.93 (7/118) | 0.435 |
| APACHE II score, median [Q1-Q3] | 22 [16-25] | 23 [16-25] | 0.219 |
| SAPS III score, median [Q1-Q3] | 58.5 [47.5-69.75] | 66 [47.5-69.75] | 0.019 |
| SOFA score, median [Q1-Q3] | 8 [6-11] | 10 [6-11] | 0.069 |
| Charlson Comorbidity Index score, median [Q1-Q3] | 1 [0-2] | 3 [0-2] | <0.001 |

Q1= quartile 1, Q3= quartile 3, APACHE= Acute Physiology and Chronic Health Evaluation, SAPS = Simplified Acute Physiologic Score, SOFA= Sequential Organ Failure Assessment.
Prepared by: authors

The median laboratory variables measured in the first 24 hours in the total group (interquartile range [Q1-Q3]) were lactate dehydrogenase 787.50 [618.25-1116.75], creatine phosphokinase 175 [88-407], procalcitonin 0.77 [0.285-2.070], leukocytes 13 810 [10 900-17 985], neutrophils 11 920 [9340-15825], lymphocytes 850 [545-1270], neutrophil/lymphocyte ratio 13.83 [8.97-22.37], D-dimers 2.22 [0.83-6.90], interleukin 6 103.40 [37.47-220.60], ferritin 1410.15 [909-2333.95], C-reactive protein 24.50 [13.20-39.90], creatinine 1 [0.78-1.60], total bilirubin 0.68 [0.44-0.97], mean platelet volume 10.4 [9.9-11.0], serum lactate 3.30 [2.20-4.40], and arterial PO_2_ 56.10 [48.75-70]. The laboratory variables between the groups of living and deceased patients are shown in Table 2.

**Table 2.** Laboratory variables in the first 24 hours.

| **Characteristics** | **Pneumonia due to SARS-CoV-2, living (n=85)** | **Pneumonia due to SARS-CoV-2, deceased (n=118)** | **P<0.05** |
| --- | --- | --- | --- |
| Lactate dehydrogenase, medium [Q1-Q3] | 787.50 [630-1119.75] | 782.50 [630-1119.75] | 0.602 |
| Creatine phosphokinase, median [Q1-Q3] | 162 [65-426] | 197 [65-426] | 0.125 |
| Median procalcitonin [Q1-Q3] | 0.63 [0.28-1.98] | 0.84 [0.28-1.98] | 0.965 |
| Leukocytes, median [Q1-Q3] | 13340 [10900-17720] | 14070 [10900-17720] | 0.849 |
| Neutrophils, median [Q1-Q3] | 11730 [9020-15150] | 12155 [9020-15150] | 0.460 |
| Lymphocytes, median [Q1-Q3] | 920 [680-1280] | 800 [680-1280] | 0.660 |
| Neutrophil/lymphocyte ratio, median [Q1-Q3] | 12.96 [8.60-18.09] | 15.69 [8.60-18.09] | 0.031 |
| D-dimers, median [Q1-Q3] | 1.40 [0.79-6.21] | 2.80 [0.79-6.21] | 0.521 |
| Interleukin 6, medium [Q1-Q3] | 94 [29.47-218.20] | 113 [29.47-218.20] | 0.654 |
| Medium ferritin [Q1-Q3] | 1408 [836.67-2331.25] | 1416.50 [836.67-2331.25] | 0.526 |
| C-reactive protein, medium [Q1-Q3] | 22.90 [12.30-38.45] | 25.40 [12.30-38.45] | 0.736 |
| Median creatinine [Q1-Q3] | 0.90 [0.70-1.30] | 1.10 [0.70-1.30] | 0.273 |
| Total bilirubin, median [Q1-Q3] | 0.66 [0.44-0.90] | 0.69 [0.44-0.90] | 0.861 |
| Mean platelet volume, median [Q1-Q3] | 10.30 [9.90-11] | 10.50 [9.90-11] | 0.762 |
| Lactate, median [Q1-Q3] | 3.00 [2.12-4.07] | 3.30 [2.12-4.07] | 0.823 |
| PO<sub>2</sub> lowest, median [Q1-Q3] | 57.80 [51-72] | 55.75 [51-72] | 0.490 |

Q1= quartile 1, Q3= quartile 3, PO<sub>2</sub> = partial pressure of oxygen
Prepared by: authors

The median variables corresponding to ventilatory support and respiratory monitoring in the first 24 hours (interquartile range [Q1-Q3]) were FiO_2_ maximum 1 [0.60-1], highest PEEP 12 [10-14], static compliance 28 [22-35.50], plateau pressure 26 [23-29], expiratory resistance 16 [13-19], highest tidal volume 400 [360-450], highest peak pressure 29 [25-31.75], lowest PaO_2_/FiO_2_ ratio 72 [55-94.50], VT/kg ratio 6.82 [6.13-7.50], and highest drive pressure 14 [11-16]. The mechanical ventilation variables between the groups of living and deceased patients are shown in Table 3.

**Table 3.** Ventilatory variables in the first 24 hours.

| **Characteristics** | **Pneumonia due to SARS-CoV-2, living (n=85)** | **Pneumonia due to SARS-CoV-2, deceased (n=118)** | **P<0.05** |
| --- | --- | --- | --- |
| FiO<sub>2</sub> maximum, median [Q1-Q3] | 1 [0.60-1] | 0.96 [0.60-1] | 0.992 |
| Highest PEEP, median [Q1-Q3] | 12 [10-14] | 12 [10-14] | 0.949 |
| Lowest static compliance, medium [Q1-Q3] | 30 [23.25-36] | 28 [23.25-36] | 0.441 |
| Plateau pressure, median [Q1-Q3] | 26 [22-29] | 26 [22-29] | 0.915 |
| Expiratory resistance, median [Q1-Q3] | 16 [13-18] | 17 [13-18] | 0.850 |
| Highest tidal volume, median [Q1-Q3] | 416 [375.25-450] | 394 [375.25-450] | 0.126 |
| Highest peak pressure, median [Q1-Q3] | 30 [25-32] | 29 [25-32] | 0.559 |
| Lowest PaO<sub>2</sub>/FiO<sub>2</sub>, median [Q1 - Q3] | 72 [56-97.50] | 70.23 [56-97.50] | 0.803 |
| VT/kg, median [Q1-Q3] | 6.88 [6.14-7.54] | 6.71 [6.14-7.54] | 0.612 |
| Highest drive pressure, medium [Q1-Q3] | 14 [11-16] | 13 [11-16] | 0.806 |

Q1= quartile 1, Q3= quartile 3, PO<sub>2</sub> = partial pressure of oxygen, FiO<sub>2</sub> = fraction of inspired oxygen, PEEP= positive end-expiratory pressure, VT= tidal volume, kg = kilograms
Prepared by: authors

All patients had median durations (interquartile range [Q1-Q3]) of 13 days of mechanical ventilation [9-20], 15 days of stay in the ICU [10-20], and 19 days of stay in the hospital [12.5-29]. Among complications, 57 patients presented with acute kidney injury (28%). Treatment included steroids in 138 patients (67.98%), vasoactive agents in 188 patients (92.61%), antimicrobials on admission in 186 patients (91.62%), IMV in the prone position in 139 (139/200) patients (69.50%), and recruitment maneuvers in 27 (27/188) patients (14.36%). The data from the groups of living and deceased patients are shown in Table 4.

**Table 4.** Variables of complications, treatments, and permanence.

| Characteristics | Pneumonia due to SARS-CoV-2, living (n=85) | Pneumonia due to SARS-CoV-2, deceased (n=118) | P<0.05 |
| --- | --- | --- | --- |
| Primary ARDS, % | 84.70 (72/85) | 94.91 (112/118) | 0.013 |
| Sepsis, % | 95.18% (79/83) | 98.30 (116/118) | 0.199 |
| Septic shock, % | 83.95% (68/81) | 85.21 (98/115) | 0.808 |
| Acute renal failure, % | 23.52% (20/85) | 31.35 (37/118) | 0.220 |
| Steroid use, % | 67.05% (57/85) | 68.64 (81/118) | 0.811 |
| Use of vasoactive agents, % | 91.76% (78/85) | 93.22 (110/118) | 0.695 |
| Antimicrobial use, % | 95.29% (81/85) | 88.98 (105/118) | 0.109 |
| Prone ventilation use, % | 63.85% (53/83) | 73.50 (86/117) | 0.144 |
| Recruitment maneuvers, % | 14.10% (11/78) | 14.54 (16/110) | 0.932 |
| Days of mechanical ventilation, median [Q1-Q3] | 13 [9-21] | 14 [10-19] | 0.675 |
| Days in ICU, median [Q1-Q3] | 16 [11-28] | 14 [11-28] | 0.067 |
| Days in hospital, median [Q1-Q3] | 27 [19-40] | 15 [19-40] | <0.001 |

Q1= quartile 1, Q3= quartile 3, ARDS= acute respiratory distress syndrome, ICU= intensive care unit. Prepared by: authors

### Primary results

In the univariate analysis, age, weight, arterial hypertension status, SAPS III score, Charlson Comorbidity Index score, the neutrophil/lymphocyte ratio, and ARDS status at admission were significantly different between the groups of living and deceased patients. A multivariate analysis was performed on the variables identified in order to identify possible risk factors for mortality, an elevated Charlson Comorbidity Index score, the presence of ARDS, and an increased neutrophil/lymphocyte ratio. The results of this analysis are presented in Table 5. In the case of the weight variable, the multivariate model demonstrated a marginal protective effect against death, with an odds ratio (OR) of 0.969 (95% confidence interval [CI]: 0.944-0.993, p = 0.015). To validate the obtained model, indicators such as the area under the ROC curve were used, whose resulting value was 75.90%, and the confusion matrix was used to determine the classification of the model in cases of mortality, resulting in 70% correct answers, as shown in Figure 1.

**Table 5:** Multivariate analysis for possible mortality risk factors.

|  | Z statistic | P value | OR | 95% CI |
| --- | --- | --- | --- | --- |
| Charlson Comorbidity Index score | 2.996 | 0.00273 | 1.365 | 1.121-1.688 |
| ARDS | 2.220 | 0.02643 | 4.165 | 1.241-16.047 |
| Neutrophil/Lymphocyte ratio | 2.195 | 0.02818 | 1.040 | 1.008-1.078 |

ARDS= acute respiratory distress syndrome, OR= odds ratio, CI= confidence interval Prepared by: authors

### Secondary results

A total of 184 patients had ARDS upon admission (90.64%, 95% CI 86.38-94.89); of these patients, 17 had mild ARDS (9.23%), 86 had moderate ARDS (46.73%) and 100 had severe ARDS (54.34%). A total of 195 (195/201) patients presented with sepsis upon admission to the ICU (97%, 95% CI 94.41-99.61), and 166 (166/196) presented septic shock (84.69%, 95% CI 79.39-89.99). A total of 118 patients died in the ICU (58%, 95% CI 51.09-65.16), and 124 patients died within 28 days of ICU admission (61%, 95% CI 54.13-68.03).

## DISCUSSION

SARS-CoV-2, which produces the disease known as “Coronavirus Disease 2019” (henceforth referred to as “COVID-19”) has been shown to produce a spectrum of diseases, ranging in severity from asymptomatic to severe cases requiring admission to the intensive care unit (ICU). A study of 72,314 cases classified 81% as mild, with no pneumonia or mild pneumonia; 14% as severe, with the presence of dyspnea, tachypnea, hypoxemia, and pulmonary infiltrates greater than 50%; and 5% as critical, with the presence of ARDS, multiorgan failure, and septic shock ^19^.

In this study, all patients were critically ill with severe CP, requiring IMV from the time of admission to the ICU; upon admission, more than 90% had a diagnosis of ARDS, and more than 85% had a diagnosis of septic shock.

The majority of cases of pneumonia caused by the novel coronavirus (SARS-CoV-2) have been observed in individuals between the ages of 20 and 59 years, with notable exceptions being Italy, Germany and Korea, where the majority of cases have been observed in individuals over the age of 65 years ^20^. Among all patients, approximately 51% are men. In Ecuador, with respect to COVID-19, data of epidemiological relevance are provided by Ortiz-Prado E et al. ^21^ from an analysis conducted between February and April 2020. The mean age of the male subjects was 42 years, and that of the female subjects was 39 years. The group aged 19-50 years was the most affected, with a fatality rate of 1.6% that increased with comorbidities and age and even varied based on ethnicity.

Our group of patients was mostly over 60 years old, possibly because the hospital in this study serves a higher percentage of older patients. There was a clear predominance of the male sex. Notably, there was a low overall Charlson Comorbidity Index score upon admission, 2 points, which would estimate a 10-year survival rate of 90% and could be explained by the fact that many patients with chronic diseases did not reach the hospital at the peak of the pandemic and possibly died at home. However, it is striking that with this score, there was a significant difference when comparing living and deceased patients; the Charlson Comorbidity Index could be considered a risk factor for death in this study.

### Risk factors

Previous studies have identified several risk factors associated with mortality, such as cardiovascular diseases, diabetes mellitus, hypertension, neoplasia, chronic lung disease, and advanced age ^20,22,23^. Other publications have mentioned advanced age ^5,7,24^, especially those over 65 years of age ^8,9^, male sex ^8^, and the presence of diabetes mellitus ^10^ as risk factors for mortality in patients with severe COVID-19. This finding was evaluated through a meta-analysis of mortality in patients with COVID-19, which revealed that mortality in COVID-19 patients is twice as high as that in nondiabetic patients. Variables that include severity upon admission to the ICU, measured by scales such as the APACHE II (an increase of 5 points) ^5^ and SOFA ^25^, are also associated with increased mortality.

Laboratory findings, such as oxygen saturation values and a PaO_2_ /FiO_2_ ratio less than 100 ^5^, in addition to several inflammatory markers upon admission and during the ICU stay, such as elevated procalcitonin, D-dimers greater than 1 µg/L ^25^, elevated lactate, a peripheral lymphocyte count (PLC) less than 0.95 × 10^9^/L ^7^ and increased platelet count ^5,9^ are also associated with a greater risk of death.

Medical complications during the evolution of the disease, described as risk factors for death, include acute renal failure, cardiac arrest, and septic shock ^5,9^, the latter of which was observed in the context of frequent nosocomial infections in this group of patients.

This study revealed that the presence of chronic diseases assessed by high Charlson Comorbidity Index score, a diagnosis of ARDS upon admission to the unit, and a high neutrophil/lymphocyte ratio (NLR) could be considered risk factors for mortality.

### Charlson Comorbidity Index

Evidence suggests that the Charlson Comorbidity Index should be used to risk stratify patients hospitalized with COVID-19, with a value ≥ 3 being associated with poor prognosis and higher mortality, and that for each point increase in the score, there is a 16% increase in the risk of death ^26^. Other reviews mention a cutoff value of > 4 adjusted for age, with an AUC of 0.709 (p = 0.001), a sensitivity of 68%, a specificity of 62% ^27^. These results coincide with the findings of our study in the group of deceased patients.

### ARDS

The mortality of ARDS as a complication of CP depends on the severity of its presentation: mild-moderate-severe, with values of 30-34-50%, respectively ^28,29^. In the case of patients with COVID-19 admitted to the ICU with ARDS, there is variability in mortality, with values of 27-89%, depending on the country ^30^; however, this risk factor would be the only one in which we could intervene and improve the prognosis, for example, by protocolizing the diagnosis and treatment of this complication. In our study, the presence of ARDS quadrupled the risk of death.

### Neutrophil/lymphocyte ratio

An elevated NLR has been proposed as a risk factor for death in critically ill patients with COVID-19. There is evidence of the usefulness of the NLR for predicting disease severity and mortality in patients with COVID-19, with area under the ROC curve values of 0.85 (95% CI 0.81-0.88) for severity and 0.90 (95% CI 0.87-0.92) for mortality ^31^. Cutoff values have been established. In a study of 2071 patients with COVID-19 admitted to the ICU, King et al. reported that a NLR greater than 7.45 increased the probability of death by 1.32 times (95% CI 1.14-1.54) ^32^. Our study has a median that doubles the aforementioned cutoff value, but the probability of death is slightly lower.

### Sepsis and ARDS

The frequency with which sepsis occurs as a secondary complication of viral infections remains uncertain. According to the findings of certain studies, 1% of patients who develop sepsis have viral infections. A prospective study from Southeast Asia published in 2018 demonstrated that viruses were the cause of up to 33% of cases of sepsis in adults. Furthermore, approximately one-third of adults requiring admission to the intensive care unit for CP have a viral infection ^33,34^.

During the pandemic caused by a SARS-CoV-2-type virus, a percentage of the population presented severe respiratory illness associated with sepsis and septic shock plus organ dysfunction ^35–37^. In this study, more than 90% of the patients had sepsis upon admission to the unit, and more than 80% met the septic shock criteria based on the SEPSIS-3 consensus.

The low procalcitonin values of less than 1 are striking, so it could be assumed that the etiology was initially viral. Importantly, the high NLR values reveal a moderate degree of physiological stress in these patients, possibly explained by viral sepsis; however, this contrasts with the high use of antimicrobials upon admission of these patients, mainly due to the lack of knowledge of this epidemic, the use of empirical treatments without further evidence and the lack of resources to establish rapid bacteriological diagnoses.

ARDS can be caused by CP due to respiratory viruses or herpes viruses. The most frequent pandemic respiratory viruses are influenza H5N1 and H1N1 2009, but ARDS can also be caused by SARS-CoV-2 ^38^. The most common clinical presentation of severe COVID-19 is acute respiratory failure compatible with ARDS ^39^, defined using the Berlin diagnostic criteria ^18^.

In patients with severe SARS-CoV-2 pneumonia, the presence of unique characteristics of ARDS has been described, especially a dissociation between the severity of hypoxemia and the parameters of its lung mechanics, with a relatively high compliance and characteristics practically never observed in most cases of ARDS ^40,41^.

In this group of patients, more than 90% met the criteria for moderate or severe ARDS upon admission to the unit, with a median PaO_2_/FiO_2 value_ of less than 100; that is, most patients had moderate-severe ARDS. All patients were managed with a protective strategy, with low VT, plateau pressures less than 30, and moderate PEEP values. Steroids were used to treat only 68% of patients, considering that at the beginning of the pandemic, their use was not protocolized, and IMV in the prone position was used in 70% of patients. In addition, unlike the group described by Gattinoni, the compliance of these patients was low from the beginning.

Before the COVID-19 pandemic, prone ventilation was a strategy used for hypoxemia refractory to other treatments in the management of patients with moderate-severe ARDS, as demonstrated by the Spanish study from Álvarez-Lerma et al., with a value of 18.2% ^42^, and the APRONET study, where the frequency of use of the prone position in patients with severe ARDS was 32.9% ^43^. There was insufficient scientific evidence for the use of corticosteroids in the management of CP before the pandemic, and there are even reports of increased mortality in patients whose pneumonia was caused by influenza. In the study by Álvarez-Lerma et al., only 40.3% of participants used corticosteroids ^42^. Since the emergence of SARS-CoV-2, the use of dexamethasone has been protocolized, as evidenced by the findings of the RECOVERY group trial ^44^. Currently, the guidelines of the American Society of Critical Care recommend the use of corticosteroids to treat ARDS patients ^45^.

### Mortality

Overall mortality in patients with CP has been estimated at 8-15% in some series ^46^, but mortality in patients with CP admitted to the ICU can reach 34-36% ^47^. These percentages can increase when pneumonia is complicated by septic shock, with values up to 25-43% ^48^, or with ARDS, with values of 30- to 50% ^28,29^.

The overall mortality of patients with COVID-19 admitted to the ICU was determined to be 30-40%; however, when mortality was analyzed by country income level, mortality among patients in ICUs in high-income countries was as low as 10.6%, and that among patients in low-income countries was as high as 79.8% ^49^. Mortality also depends on geography; in the study published by Qian et al., mortality in 2020 was 39% in China, 48% in the rest of Asia, 34% in Europe, 15% in the United States, and 39% in Eastern Europe ^50^. In Latin America, the reported mortality of critically ill patients with COVID-19 receiving IMV was 57%, reaching up to 63.3% in some countries ^51–53^. The ICU mortality in this study was similar to that reported in low-income countries for patients with severe ARDS and septic shock.

### Limitations

This was a single-center, retrospective study that did not include patients with more than 48 hours of clinical management before admission to the ICU due to variability in management outside the unit. The frequency of microbial superinfection upon admission or during the ICU stay, a factor that could influence mortality and was limited by the exhaustion of resources, was not reported in this study.

## CONCLUSIONS

An elevated Charlson Comorbidity Index score, the presence of ARDS, and an increased NLR upon admission in critically ill patients with severe CP due to SARS-CoV-2 receiving IMV could be risk factors for mortality. The frequency of ARDS and sepsis upon admission in this group of patients was high, and mortality in this study was within the values reported for Latin American countries, which have low to moderate economic incomes, for patients with severe ARDS and septic shock.

## Data Availability

All data produced in the present study are available upon reasonable request to the author

## Acknowledgements

Our thanks to all the health personnel of the Intensive Care Unit.

